# High incidence of Y-chromosome mosaicism in male and female individuals with MOGHE

**DOI:** 10.1101/2025.10.15.25337897

**Authors:** Erica Cecchini, Till Hartlieb, Ahmed Gaballa, Katja Kobow, Mitali Katoch, Georgia Vasileiou, Wiebke Hofer, Marie Reisch, Manfred Kudernatsch, Christian G. Bien, Roland Coras, Ingmar Blümcke, Lucas Hoffmann

**Affiliations:** Department of Neuropathology, Universitätsklinikum Erlangen, Friedrich-Alexander-Universität (FAU) Erlangen-Nürnberg, Erlangen, Germany, and partner of the European Reference Network (ERN) EpiCARE; Centre for Paediatric Neurology, Neurorehabilitation, and Epileptology, Schoen-Clinic, Vogtareuth, Germany; Research Institute for Rehabilitation, Transition, and Palliation, Paracelsus Medical University, Salzburg, Austria; Department of Epileptology, Krankenhaus Mara, Bethel Epilepsy Centre, Medical School OWL, Bielefeld University, Bielefeld, Germany; Institute of Human Genetics, Friedrich-Alexander-Universität Erlangen-Nürnberg, Erlangen, Germany; Centre for Rare Diseases Erlangen (ZSEER), Universitätsklinikum Erlangen, Erlangen, Germany

**Keywords:** Epilepsy, SLC35A2, Somatic, CNV, oligodendroglia

## Abstract

Mild malformation of cortical development with oligodendroglial hyperplasia in epilepsy (MOGHE) is a recently discovered histopathological lesion entity. Approximately half of affected individuals carry a pathogenic brain mosaicism in the X-linked *SLC35A2* gene, and all suffer from epilepsy. In this work, we extended the search for genetic alterations of MOGHE by investigating sex chromosome copy number alterations in 29 brain tissue samples from 19 males and 10 females with histopathologically confirmed MOGHE. Twenty individuals carried pathogenic *SLC35A2* variants, while no genetic alteration was identified in nine individuals using targeted deep panel sequencing. Interestingly, DNA methylation-derived copy number variation (CNV) plots revealed significant gains of the Y chromosome in 16/19 males (84.2%) and in 5/10 females (50%). These findings were validated by chromogenic and fluorescent *in situ* hybridisation (ISH), PCR amplification of Y-specific sequences, and microscopic localisation of cells with Y-chromosomal gain in clusters of oligodendroglial hyperplasia. PCR and ISH demonstrated lesion-restricted Y-chromosome gains, absent in the overlying non-lesional neocortex. Together with pathogenic variants in the X-chromosomal *SLC35A2* gene, Y-chromosomal sequences detected in phenotypic females and mosaic Y chromosome gains in males provide a genomic correlate for all cases of MOGHE. Based on *SLC35A2* mutational status and Y-chromosome copy number changes, we stratified the cohort into three subgroups: *SLC35A2*-mutant without Y gain (**SLC+/Y–,** *n* = 8), *SLC35A2*-mutant with Y gain (**SLC+/Y+,** *n* = 12), and *SLC35A2*-wild type with Y gain (**SLC–/Y+,** *n* = 9). These genetically defined subgroups also differed in their clinical presentation, with individuals from group 2 having the earliest disease onset and the largest lesion volume on MRI. These findings expand the genetic spectrum of epileptogenic cortical malformations and highlight a potentially overlooked role of sex chromosome biology in this focal epilepsy.

## Introduction

Mild Malformation of Cortical Development with Oligodendroglial Hyperplasia in Epilepsy (MOGHE) is an increasingly recognised clinicopathological entity associated with drug-resistant epilepsy. The lesion is frequently detected in young children and localised to the frontal lobe. Surgical resection is the main therapeutic intervention for seizure control ^1,2^. First described by the Erlangen Neuropathology team in 2017^3^, MOGHE was included in the International League Against Epilepsy (ILAE) Classification of Focal Cortical Dysplasia in 2022^4^. The disease is histopathologically defined by focal areas of increased oligodendroglial cell density and increased numbers of heterotopic neurons within the white matter. Recent studies have described hypomyelinated regions within the lesion ^1,5,6^, corresponding to areas of oligodendroglial hyperplasia ^6^. Patients with MOGHE present with two different clinical phenotypes. The first one is early-onset epileptic encephalopathy (EE), which manifests with epileptic spasms and is frequently associated with significant cognitive impairment. The second phenotype is drug-resistant focal epilepsy (DR-FE), with onset typically in adolescence or young adulthood and accompanied by normal or borderline cognitive function ^1^. In the 2022 ILAE classification, MOGHE is defined as a white matter abnormality, identifiable through MRI ^7^. Neuroradiologically, lesions show mild cortical thickening, cortical/subcortical hyperintense T2/fluid-attenuated inversion recovery signal associated with grey/white matter blurring or reduced cortico-medullary differentiation in older individuals ^7,8^.

Deep-targeted gene sequencing of surgically resected brain tissue using MOGHE from multicentre paediatric cohorts has provided insights into the molecular pathogenesis of the disease condition ^5,1,9,10^. These studies have consistently identified pathogenic variants in the X-linked *SLC35A2* gene in ∼50% of individuals with MOGHE ^5,6^. Germline *de novo* heterozygous and a few hemizygous pathogenic variants in this gene, some of the latter occurring in a mosaic state, have been associated with the X-linked congenital disorder of glycosylation type IIm (MIM 300896). Affected individuals typically present with infantile-onset seizures, severe developmental delay and intellectual disability, skeletal anomalies, as well as brain malformations, including cerebellar atrophy and white matter abnormalities^11^. *SLC35A2* encodes a UDP-galactose transporter responsible for moving galactose from the cytosol into the lumen of the Golgi apparatus, a process essential for the glycosylation of proteins and lipids. Multiple studies have demonstrated a consistent association between *SLC35A2* variants and MOGHE^5,6^, as well as with intractable neocortical epilepsy^12^, supporting its role in disease pathogenesis. Recent advances in animal modelling have provided further understanding of the pathogenic role of *SLC35A2* in cortical development. Multiple experimental approaches have shown that loss of *SLC35A2* disrupts corticogenesis by impairing neuronal migration and altering neuronal excitability, while also inducing oligodendroglial hyperplasia and hypomyelination ^13–15^. Furthermore, recent evidence has demonstrated a significant reduction of SLC35A2 protein in the brain white matter of individuals harbouring *SLC35A2* nonsense variants ^6^, suggesting that the nature of the genetic alteration may influence therapeutic response. Specifically, interindividual variability of *SLC35A2* variants may underlie differential responses to D-galactose supplementation, with some patients exhibiting clinical benefit while others remain refractory ^16^. Nevertheless, in roughly half of cases, no pathogenic variant in *SLC35A2* or other established lesional epilepsy genes has been detected, leaving their genetic aetiology unexplained^5,9,17,18^.

To date, genetic epilepsies have been primarily attributed to Single Nucleotide Variants (SNVs) in autosomal or X-linked genes ^19^. Although sex chromosome aneuploidies are increasingly recognised as significant contributors to neurodevelopmental disorders and broader neurological impairment ^20–25^, their relationship with epilepsy remains less clearly defined. Nonetheless, seizures have been consistently reported in individuals with sex chromosome disorders such as Turner syndrome [45, X]^26,27,28–30^, Klinefelter syndrome [47, XXY]^31–34^, 47, XYY syndrome [47, XYY]^35–37^, and 47, XXX syndrome [47, XXX]^38–40^. While most of the literature has emphasised X chromosome aberrations as sex chromosomal contributors to neurological dysfunction and epilepsy, the epileptogenic potential of Y chromosome aneuploidies and mosaicism has only rarely been considered. Notably, case reports and small series describe epilepsy or electroencephalographic abnormalities in a subset of individuals with 47, XYY syndrome ^37,41^, and population-based studies suggest increased neurological morbidity in this group ^42–45^. Y-chromosome mosaicism [45, X/46, XY] or cryptic Y-material can be detected in blood or other tissues in 5-12% of females with Turner syndrome ^46,47^. Such cases illustrated that Y-sequence mosaicism can occur in individuals without overt male phenotype and may be tissue-restricted.

Given these observations, it is plausible that structural abnormalities or mosaicism involving the Y chromosome could contribute to epileptogenesis. To date, however, no studies have systematically investigated Y chromosome gain, loss, or mosaicism in MOGHE. In this context, our study demonstrates recurrent Y chromosome gains in patients with MOGHE, detected selectively in lesional white matter but absent from adjacent non-lesional cortex. This intra-sample comparison provides evidence for lesion-restricted Y chromosome gain mosaicism or microchimerism as a potential pathogenic mechanism in MOGHE.

## Materials and methods

### Cohort Selection

Brain tissue samples were retrieved from the archive of the Neuropathological Institute of Erlangen University Hospital, the Bethel Epilepsy Centre, and the Schoen Clinic Vogtareuth in Germany. All samples were histopathologically evaluated for the presence of MOGHE by two independent board-certified neuropathologists (Ingmar Blümcke and Roland Coras) according to the most recent International League Against Epilepsy (ILAE) diagnostic guidelines^4^. Diagnostic criteria included oligodendroglial hyperplasia, defined as increased density of oligodendroglial cells in subcortical white matter, focal myelin loss confirmed by Nissl-LFB staining, and the presence of heterotopic neurons within the white matter. The cohort consisted of 29 individuals (19 males and 10 females) with histologically confirmed MOGHE. Clinical metadata, including sex, age range at epilepsy onset, epilepsy phenotype, MRI findings, and surgical outcome, were collected from patient records and are summarised in Table 1. Tissue specimens were formalin-fixed and paraffin-embedded (FFPE) for histopathology and molecular studies. The study was approved by the ethics committees of the Medical Faculty of the Friedrich-Alexander University of Erlangen-Nuremberg (#193_18B, 18-192_1-Bio) and the University of Münster (2015-088-f-S). Written informed consent was obtained from all patients or their legal guardians.

**Table 1.** Patients’ clinical and molecular data. **Legend Table 1:**Abbreviations: F: female; M: male; R: right; L: left; F: frontal; T: temporal; Fb: fronto-basal; Fp: fronto-polar; Fm: fronto-mesial; Fop: fronto-orbito-polar; Fl: fronto-lateral; Gy: gyrus; Hm: hemispheric; TPO: Temporo-Parieto-Occipital. NPSY: Neuropsychology; refers to the neuropsychological assessment before and after surgery. N.s.: non-significant; n.a.: non-available. **\*** Confirmed by PCR analysis. The CNV profile for this patient was inconclusive and was subsequently validated by FISH and PCR. **^b^** The CNV profile for this patient was inconclusive and was subsequently validated by FISH.

| ID | Sex | Lesion Size | SLC35A2 Variant (hg19)<br>NM_005660.3 (cDNA change) | NP_005651.1 (amino acid change) | SLC35A2 Variant type | VAF | Y-gain (CNV log2 ratio) | Outcome (Engel) | NPSY (Pre → Post) |
| --- | --- | --- | --- | --- | --- | --- | --- | --- | --- |
| **Group 1: SLC+/Y-, (n = 8)** |  |  |  |  |  |  |  |  |  |
| 1. | M | F-R | c.533G>T | p.(Gly178Val) | missense | 26,9% | No | 1A | Mild/moderate → Mild/moderate |
| 2. | M | Fb-R | c.335_339dup | p.(Lys114ArgfsTer32) | frameshift | 52% | No | 1A | Borderline → Average |
| 3. | M | F-R | c.206C>T | p.(Thr69Leu) | missense | 14% | No | 1A | Severe → Severe |
| 4. | F | Fm-R | c.935C>T | p.(Ser312Phe) | missense | 8% | No * | 1A | Borderline → Mild |
| 5. | F | F-L | c.905C>T | p.(Ser302Phe) | missense | 8% | No * | 4A | Severe → Severe |
| 6. | F | Fop-R | c.640G>C | p.(Gly214Arg) | missense | 8% | No | 4B | No intellectual impairment |
| 7. | F | FI-R | c.515T>C | p.(Leu172Pro) | missense | 3% | No | 3A | No intellectual impairment |
| 8. | F | F Gy-R | c.364_365insC<br>c.366delinsTCTC | p.(Tyr122SerfsTer6)<br>p.(Tyr122_Thr123insLeu) | frameshift<br>in frame | 15% | No | 3A | No estimate |
| **Group 2: SLC+/Y+, (n = 12)** |  |  |  |  |  |  |  |  |  |
| 9. | F | Hm-L | c.359_360delTC | p.(Leu120HisfsTer7) | nonsense | 30% | Yes <sup>a,*</sup> | 4A | n.a. |
| 10. | F | Hm-L | c.603_606dupAGGC | p.(Leu203ArgfsTer20) | frameshift | 21% | Yes <sup>a,*</sup> | 1A | Severe → Moderate |
| 11. | F | F-R | c.640G>C | p.(Gly214Arg) | missense | n.a. | Yes (0,0) * | 1A | n.a. |
| 12. | F | Hm-L | c.935C>T | p.(Ser312Phe) | missense | 33% | Yes (0,3) * | 1A | n.a. |
| 13. | M | Hm-L | c.424C>T | p.(Gln142X) | nonsense | 19,4% | Yes (0,4) | 1A | Severe → Severe |
| 14. | M | Hm-R | c.569_572del | p.(Gly190AlafsTer158) | frameshift | 41% | Yes<sup>b</sup> | 3A | Severe → Severe |
| 15. | M | TPO-L | c.832C>T | p.(Gln278Ter) | nonsense | 48% | Yes (0,9) | 1A | Severe → Severe |
| 16. | M | FT-R | c.675dupA | p.(Gly226ArgfsTer29) | nonsense | 40% | Yes (0,9) | 1A | Moderate → Borderline |
| 17. | M | F-L | c.113delT | p.(Ile38AsnfsTer3) | frameshift | 41% | Yes (0,9) | 1A | Moderate → Moderate |
| 18. | M | FT-L | c.580_616dup | p.(Val206AlafsTer28) | nonsense | 9% | Yes (1,0) | 3A | n.a. |
| 19. | M | F-L | c.665_667del | p.(Lys222del) | in frame | 32,6% | Yes (0,8) | 3A | n.a. |
| 20. | M | Hm-L | c.322C>T | p.(Gln108Ter) | missense | 25% | Yes (1,0) | 1A | Severe → Moderate |
| **Group 3: SLC-/Y+, (n = 9)** |  |  |  |  |  |  |  |  |  |
| 21. | F | F-R | wt | - | No Var. | - | Yes (1,0)<br>* | n.a. | n.a. |
| 22. | M | F-L | wt | - | No Var. | - | Yes (0,4) | 1A | Severe → n.a. |
| 23. | M | Fp/Fb-L | wt | - | No var. | - | Yes (0,4) | 1A | Moderate → Moderate |
| 24. | M | TPO-L | wt | - | No Var. | - | Yes (0,4) | 4A | Severe → Severe |
| 25. | M | Fb-L | wt | - | No Var. | - | Yes (0,8) | 1A | No intellectual impairment |
| 26. | M | T G-R | wt | - | No Var. | - | Yes (0,8) | n.a. | n.a. |
| 27. | M | F-R | wt | - | No Var. | - | Yes (0,9) | 2A | Borderline → Borderline |
| 28. | M | F-R | wt | - | No Var. | - | Yes (1,0) | 1A | Moderate → Mild |
| 29. | M | Hm-L | wt | - | No Var. | - | Yes (1,0) | 1A | n.a. |

### Genetic background of the cohort

All of the 29 MOGHE patients included in this study had previously been reported, 20 of them carrying pathogenic variants in the X-linked *SLC35A2* gene (68.9%) ^1,6,9,17,48^. In these earlier studies, targeted deep sequencing of resected brain tissue identified ten patients with truncating variants and ten with missense substitutions, with variant allele frequencies (VAF) ranging from 3% to 52%. The remaining nine patients did not harbour pathogenic variants in *SLC35A2* or any other epilepsy-related gene. Because these genetic data have already been published, we did not repeat or expand sequencing analyses in the present study. No matched blood samples were available to test for potential germline contribution; however, the published evidence supports the somatic nature of the mutations identified in brain tissue.

### Neuropsychological Assessment

At the Schoen Clinic Vogtareuth, intellectual functioning was evaluated a few days prior to epilepsy surgery and at 6 and 24 months postoperatively in 13 of 17 patients residing in German-speaking countries. Standardised, age-appropriate instruments were applied, including the Hamburg-Wechsler-Intelligenztest für Kinder III (HAWIK-III; Tewes et al., 1999), the Wechsler-Intelligenztest für Erwachsene (WIE; von Aster et al., 2006), the Bayley Scales of Infant Development II (BSID-II; Bayley, 1993), and the Vineland Adaptive Behaviour Scales (VABS; Sparrow et al., 1984). Intellectual functioning was classified according to ICD-10 criteria for intellectual disabilities (F70–F73): (1) no impairment (IQ ≥ 85), (2) borderline (IQ 70–84), (3) mild (F70; IQ 50–69), (4) moderate (F71; IQ 35–49), and (5) severe/profound (F72/73; IQ < 35).

At the Bethel Epilepsy Centre, neuropsychological assessment included full-scale IQ testing in children preoperatively and at 6 (PO6) and 24 months (PO24) after surgery. The instruments used were the Wechsler Intelligence Scale for Children (WISC-IV; Petermann & Petermann, 2011; WISC-V; Petermann, 2017), the Wechsler Preschool and Primary Scale of Intelligence (WPPSI-III; Petermann et al., 2014; WPPSI-IV; Petermann & Daseking, 2018), and the McCarthy Scales of Children’s Abilities (MSCA; McCarthy, 1972). In adults, general intellectual ability was estimated preoperatively using a vocabulary test (Mehrfachwahl-Wortschatz-Intelligenztest, MWT-B; Lehrl, 1989) and a logical reasoning test (Leistungsprüfsystem 4, LPS-4; Horn, 1962).

### DNA Methylation and CNV Analysis

DNA methylation was profiled using the Illumina Infinium MethylationEPIC BeadChip (850k) array. Raw IDAT files were processed and normalised with the minfi R package (v1.46.0) using Noob normalisation, and mapped to the hg19 genome build. CNVs were inferred from methylation intensity data using a customised extension of the conumee R package [github.com/FAU-DLM/conumee.git], originally forked from the standard conumee, which incorporates enhanced summary-plot functionality for improved CNV visualisation. Normalisation was performed against in-house control samples. Gene annotations were obtained from EnsDb.Hsapiens. v86 and restricted to protein-coding genes on standard UCSC chromosomes. Probes were further filtered against the Illumina Human Methylation

EPICanno.ilm10b4.hg19 annotation to retain high-confidence loci. Significant CNV gains of Y chromosome–associated sequences were defined as log2 ratios >0.4 for males and >0.0 for females. Quality control included filtering for minimum probe coverage per bin, exclusion of low-confidence regions, and use of matched controls for reference normalisation. Summary-level and sample-specific CNV plots were generated across autosomes and sex chromosomes.

### Chromogenic and Fluorescent *in Situ* Hybridisation

Chromogenic *in situ* hybridisation (CISH) was performed using the ZytoDot2C CISH Implementation Kit and the corresponding CEN X/Y probes (ZytoVision, Germany), following the manufacturer’s protocol with minor modifications to the digestion time optimised for brain tissue. The ZytoDot 2C CEN X probe consists of dinitrophenyl-labelled polynucleotides targeting sequences within Xp11.1-q11.1, specific for the centromeric region of chromosome X, detected in red. The ZytoDot 2C CEN Y probe comprises digoxigenin-labelled polynucleotides targeting sequences within Yp11.1-q11.1, specific for the centromeric region of chromosome Y, detected in green. Fluorescence *in situ* hybridisation (FISH) was carried out using the ZytoLight FISH-Tissue Implementation Kit with the CEN X/Y dual-colour probe (ZytoVision) for chromosome enumeration. The dual-colour probe includes ZyGreen-labelled polynucleotides (excitation 503 nm, emission 528 nm) targeting the centromeric region of chromosome X (Xp11.1-q11.1), and ZyOrange-labelled polynucleotides (excitation 547 nm, emission 572 nm) targeting the centromeric region of chromosome Y (Yp11.1-q11.1, alpha satellite DNA). As additional controls for FISH, one male individual with temporal lobe epilepsy (TLE; aged 0–5 years) and one female individual with TLE (aged 10–15 years) were included to confirm the expected X/Y signal patterns in non-MOGHE brain tissue.

FFPE tissue sections were first incubated at 70°C for 30 minutes to facilitate paraffin removal, followed by deparaffinization in xylene and rehydration through a graded ethanol series. Endogenous peroxidase activity was quenched with 3% hydrogen peroxide for 5 minutes, and heat-induced epitope retrieval was performed using a pre-warmed EDTA buffer (PT2) at 95°C for 15 minutes. Proteolytic digestion was conducted with Pepsin at 37°C for 7 minutes. After dehydration and air-drying, probe hybridisation was initiated by applying the probe, sealing the coverslip with Fixogum, denaturing at 74°C for 5 minutes, and incubating overnight at 37°C in a humid chamber. On the second day, post-hybridisation washes were carried out, and probe detection was achieved using anti-DIG/DNP antibody mixes, HRP/AP-Polymer-Mix, and chromogenic substrates (AP-Red and HRP-Green). Nuclear counterstaining was performed with Nuclear Blue, followed by dehydration, xylene clearing, and mounting.

CISH slides were digitised with a Hamamatsu S60 scanner (Japan). FISH slides were examined at 40x/0.75 magnification using an Olympus BX51 Fluorescence Microscope (Japan), and images were captured with an Olympus ColorView 2 camera with Soft Imaging System via a filter cube designed for FISH. Although the Y-probe is labelled with ZyOrange, we visualised it as red due to filter limitations; accordingly, figures display the Y chromosome signals in red. The distribution of chromosome-specific signals within cortical and subcortical areas were recorded.

### PCR Amplification of Y-Specific Sequences

PCR amplification of the sex-determining region Y (*SRY*) locus was performed as a molecular confirmation step. Manual microdissection of white matter (lesion) and overlying cortex (non-lesion) within the same FFPE block was performed, enabling intra-sample comparison.

Genomic DNA was extracted using the QIAamp DNA Mini Kit (Qiagen), optimised for micro-dissected tissue. Due to the limited DNA yield from MOGHE brain tissue and the small size of the dissected samples (restricted to the lesion site of oligodendroglial clusters), a two-step PCR amplification protocol was implemented. The first step consisted of 35 cycles of amplification using 15µl containing 20ng/µl of DNA. Thermal cycling parameters were as follows: initial activation at 95 °C for 5 minutes, denaturation at 95 °C for 30 seconds, annealing at 64 °C for 90 seconds, and extension at 72 °C for 90 seconds, followed by a final extension at 68 °C for 10 minutes. A secondary amplification was then performed using a 1:15 dilution of the primary PCR product to ensure sufficient yield for downstream analysis. PCR primers targeting a 266 bp fragment of the sex-determining region Y (SRY) locus were purchased from Invitrogen and had the following sequences: forward 5’-GGA AGC AAA CTG CAA TTC TTC GG-3’ and reverse 5’-GAT AGA GTTG AAG CGA CCC ATG AA-3’.

PCR products were analysed on 3% agarose gels stained with Red Safe Nucleic Acid Staining Solution (Intron Biotechnology Inc.), and bands were visualised under UV light. Negative controls (genomic DNA from females with and without MOGHE) and positive controls (genomic DNA from males with and without MOGHE) were included in all runs.

Manual microdissection was performed to separately isolate cortical and white matter regions from the same FFPE tissue block. Qualitative PCR validation for Y-positive females was performed on 7 female cases.

## Results

### DNA methylation-derived CNV analysis reveals Y chromosome gains

CNV analysis based on DNA methylation profiles was performed on 29 MOGHE patients (19 males, 10 females). Significant gains of Y chromosome-associated sequences were detected in 16 of 19 male patients (84.2%) with log2 ratios ranging from +0.4 to +1.2, exceeding the pre-defined thresholds for CNV calling (**Fig. 1A**). Unexpectedly, Y chromosome gains were also detected in 5 of 10 female patients (50%), with log2 ratios ranging from +0.0 to +1.0. (**Fig.1B**). In two female patients (ID 9 and ID 10), CNV profiles were inconclusive, and the presence of Y-chromosomal material was subsequently confirmed by FISH and PCR. Similarly, in one male patient (ID 14), the CNV plot was inconclusive and was validated by FISH. No significant copy number alterations were detected on autosomes or the X chromosome across the whole cohort. The individual CNV plots for all patients are shown in Supplementary Data (**Supplementary** Fig. 1).

**Figure 1.**
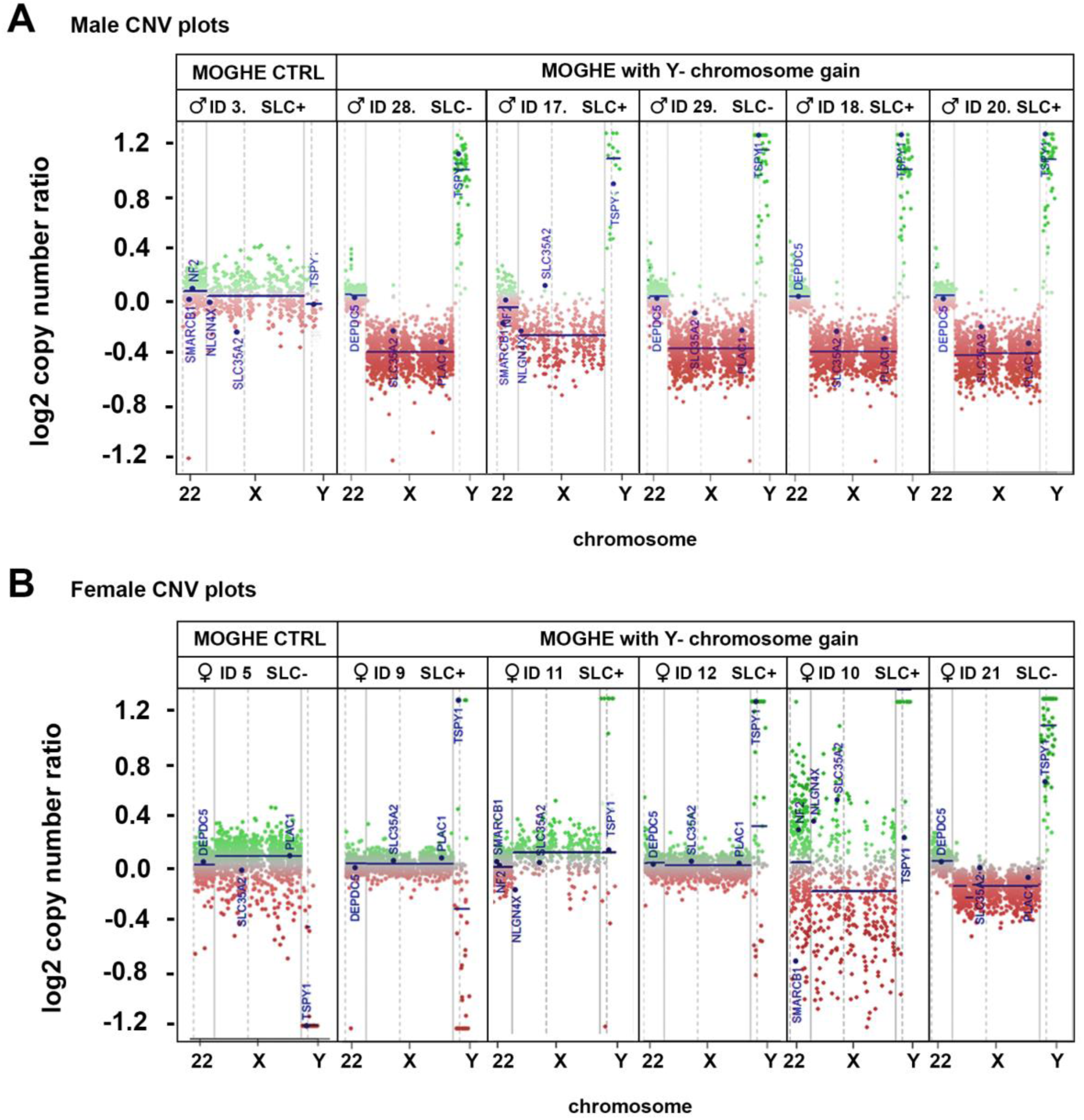
DNA methylation–derived CNV plots reveal Y chromosome gains in male and female patients with MOGHE. **Legend Figure 1.** For both panels, the x-axis indicates chromosomes 22, X, and Y, providing spatial reference within the CNV plots. The y-axis represents log₂ copy number ratios, with the baseline corresponding to the blue line at log₂ = 0.0. Annotations denote the mutational status of *SLC35A2*: “**SLC+**^+^” indicates a pathogenic variant, whereas “**SLC-**” indicates wild type. **(A)** CNV profiles of six male patients with MOGHE (ID 3, ID 28, ID 17, ID 29, ID 18, ID 20). The first case shows a normal CNV pattern (Y chromosome at log₂ ratio = 0.0), whereas the remaining five cases exhibit a gain of the Y chromosome, consistently detected between 0.8 and 1.0 on the log₂ copy number ratio scale (highlighted in green). (**B**) CNV profiles of six female patients with MOGHE (ID 5, ID 9, ID 11, ID 12, ID 10, ID 21). The first case shows no Y chromosome gain, while the remaining five cases demonstrate Y chromosome gains ranging from 0.0 to 1.0 on the log₂ copy number ratio scale. ID 9 exhibits an intermediate CNV profile that is challenging to interpret, and ID 10 displays low-quality CNV data, necessitating validation by orthogonal methods (FISH, PCR).

Based on the integrated analysis of *SLC35A2* mutational status and Y chromosome CNVs, the cohort was stratified into three distinct subgroups. **Group 1**: patients harbouring pathogenic *SLC35A2* variants without detectable Y chromosome gains (**SLC+/Y–,** *n* = 8), **Group 2**: patients carrying pathogenic *SLC35A2* variants in conjunction with Y chromosome gains (**SLC+/Y+,** *n* = 12), and **Group 3**: patients lacking *SLC35A2* mutations but exhibiting Y chromosome gains (**SLC–/Y+,** *n* = 9), as reported in Table 1.

### Clinical findings in genetically defined MOGHE subgroups

Median age ranges at seizure onset differed across the three molecularly defined subgroups. Patients with Y chromosome gains tended to present earlier than those without, although this difference did not reach statistical significance (Kruskal–Wallis test, H = 2.786, p = 0.248). Median onset ages (interquartile range, IQR, which represents the middle 50% of the data) were 3.88 years (IQR 0.54–13.5; range 0.16–24 years) for Group 1 (SLC+/Y–), 0.58 years (IQR 0.455–1.04; range 0.25–3 years) for Group 2 (SLC+/Y+), and 2.0 years (IQR 0.54–5.25; range 0.33–12 years) for Group 3 (SLC–/Y+).

Lesion size and distribution also differed across the three subgroups. Patients in Group 1 typically harboured smaller, more circumscribed lesions confined to the frontal lobe, including frontobasal, frontomesial, and orbitopolar regions (**Fig. 2A**). By contrast, Group 2 exhibited a markedly larger lesion burden, with half of the patients (6/12) showing hemispheric involvement and three patients displaying lesions spanning two or more lobes (**Fig. 2C**). Group 3 demonstrated an intermediate-to-large lesion extent, including one patient with hemispheric disease, one with the involvement of the Temporo-Parieto-Occipital lobes, and the majority with lesions encompassing the entire frontal lobe (**Fig. 2B**). When correlated with age ranges at onset, these data suggest a trend in which more extensive lesions are associated with earlier seizure manifestation, particularly in Group 2, which combined the largest lesion size with the earliest median onset.

**Figure 2.**
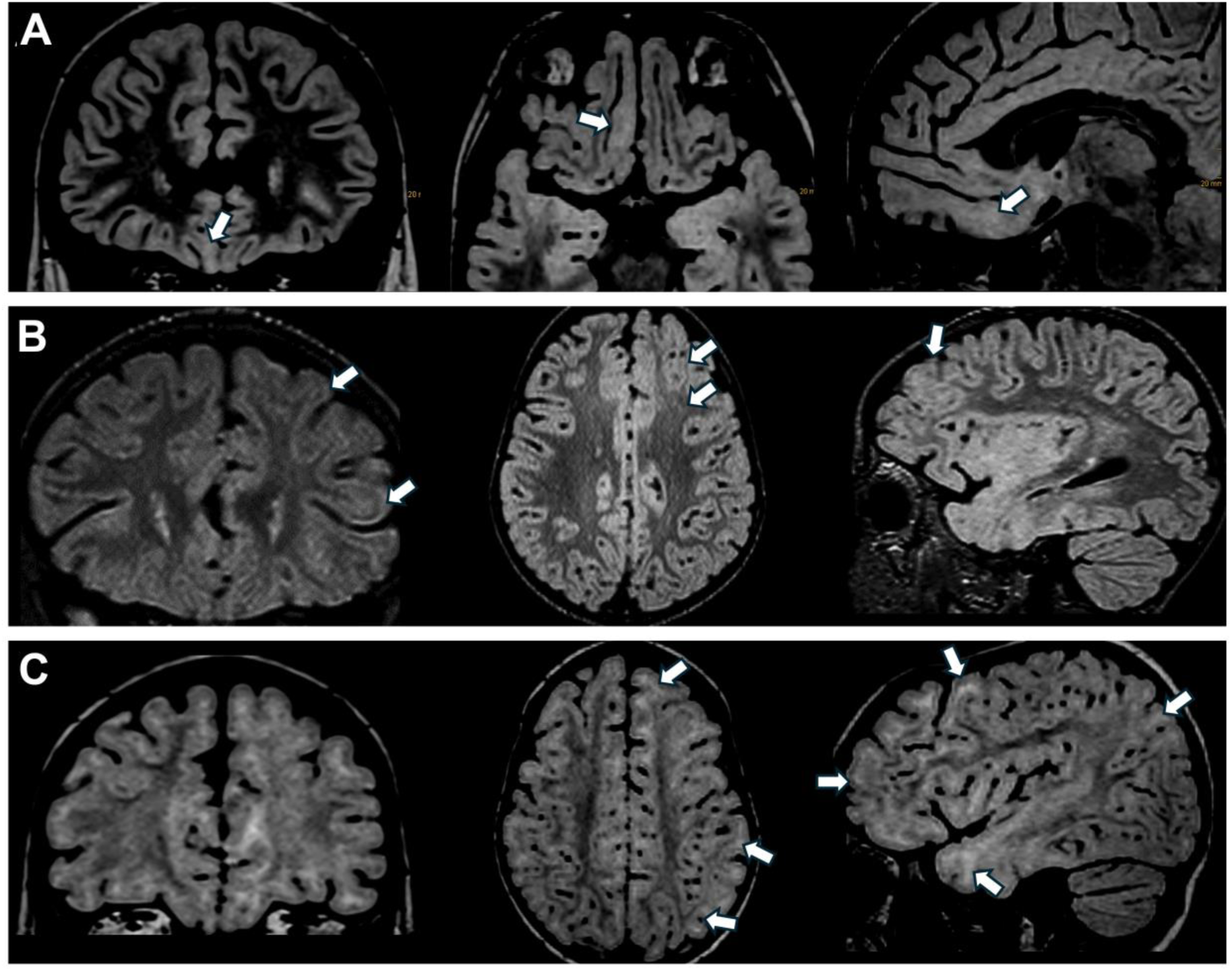
MRI images of representative MOGHE lesions. **Legend to Figure 2.** Representative MRI images in coronal, axial, and sagittal planes illustrating lesion extent and distribution. White arrows indicate the lesions. (A) Circumscribed right fronto-basal lesion. (B) Left frontal lesion. (C) Extensive left hemispheric lesion.

Pre- and postoperative neuropsychological assessments were available in 16 individuals across the three molecularly defined MOGHE subgroups. In **Group 1** (SLC+/Y–), intellectual functioning ranged from severe to average, with two patients showing postoperative improvement (borderline→average; borderline→mild) and two patients with no intellectual impairment, while the remaining cases remained stable. In **Group 2** (SLC+/Y+), cognitive impairment was generally more pronounced, with most individuals exhibiting severe or moderate intellectual disability both before and after surgery. Isolated cases demonstrated improvement (severe→moderate; moderate→borderline). In **Group 3** (SLC–/Y+), intellectual functioning was heterogeneous, spanning from severe intellectual disability to normal cognition, with most patients showing stable trajectories and a few mild postoperative gains (moderate→mild). Overall, neuropsychological outcomes across all three subgroups indicated stability or modest postoperative improvement in intellectual functioning, with no evidence of cognitive decline. Among the subgroups, **Group 1** (SLC+/Y–) exhibited the most favorable cognitive profile, **Group 2** (SLC+/Y+) the most severe impairments, and **Group 3** (SLC–/Y+) the broadest variability in outcome.

Histopathological examination revealed no significant differences among the three molecularly defined MOGHE subgroups. Quantitative assessment of oligodendroglial density (OLIG2-positive nuclei per mm² of white matter) yielded comparable mean values across groups (**Group 1: SLC+/Y-**: 2416 cells/mm²; **Group 2: SLC+/Y+**: 2376 cells/mm²; **Group 3: SLC–/Y+**: 2153 cells/mm²; *p* > 0.05; counts based on at least four representative cases per group). Proliferative activity was uniformly low, with Ki-67 indices below 1% in all samples. The extent of hypomyelinated areas on Nissl–Luxol Fast Blue (LFB) staining differed slightly between groups, being least prominent in the **SLC+/Y-**subgroup, present in 7/12 cases of the **SLC+/Y+** group, and most frequent in the **SLC–/Y+** group. This pattern aligns with our previous observations that hypomyelination decreases with patient age, such that patchy areas are more evident in younger individuals and tend to diminish in older patients ^6^.

### In-Situ Hybridisation analyses confirmed the presence of extra-numerary Y-chromosomes in a subset of individuals with MOGHE

To validate the CNV findings at the cellular level, we performed fluorescence in situ hybridisation (FISH) and chromogenic in situ hybridisation (CISH) using dual-colour centromeric X and Y probes (ZytoVision). FISH provided a clear visualisation of Y-positive cells, predominantly localised to areas of oligodendroglial hyperplasia (**Fig. 3**), with the X chromosome labelled in green, and the Y chromosome labelled in red. In female individuals with MOGHE exhibiting Y-chromosome gains in CNV analysis, including the individual with intermediate CNV results (ID 9), FISH detected Y-positive nuclei in all cases, thereby confirming the presence of Y-chromosomal sequences even when CNV plots were inconclusive. Scattered nuclei in lesional white matter displayed an additional Y signal in otherwise XX cells. In contrast, female individuals without CNV-detected Y gain showed only the two expected X signals, serving as an internal negative control. Similarly, male individuals with MOGHE with CNV-identified Y-Y-chromosome gains exhibited nuclei containing one X signal and two Y signals, confirming the presence of extra Y chromosomal material. CISH was used as an orthogonal validation method and qualitatively supported the FISH observations, with the X chromosome visualised in red and the Y chromosome in green (**Fig. 4**); however, fluorescence imaging provided higher resolution and clarity. Due to technical limitations, including the frequent truncation of nuclei in tissue sections that prevents consistent visualisation of the Y probe signal, quantitative assessment of nuclear counts was not feasible; instead, evaluation was based on qualitative assessment of signal distribution. Importantly, Y-positive nuclei were confined to lesional white matter and were absent from overlying cortex and CNV-negative controls, indicating that Y-chromosome gains are spatially restricted and associated with areas of oligodendroglial hyperplasia.

**Figure 3.**
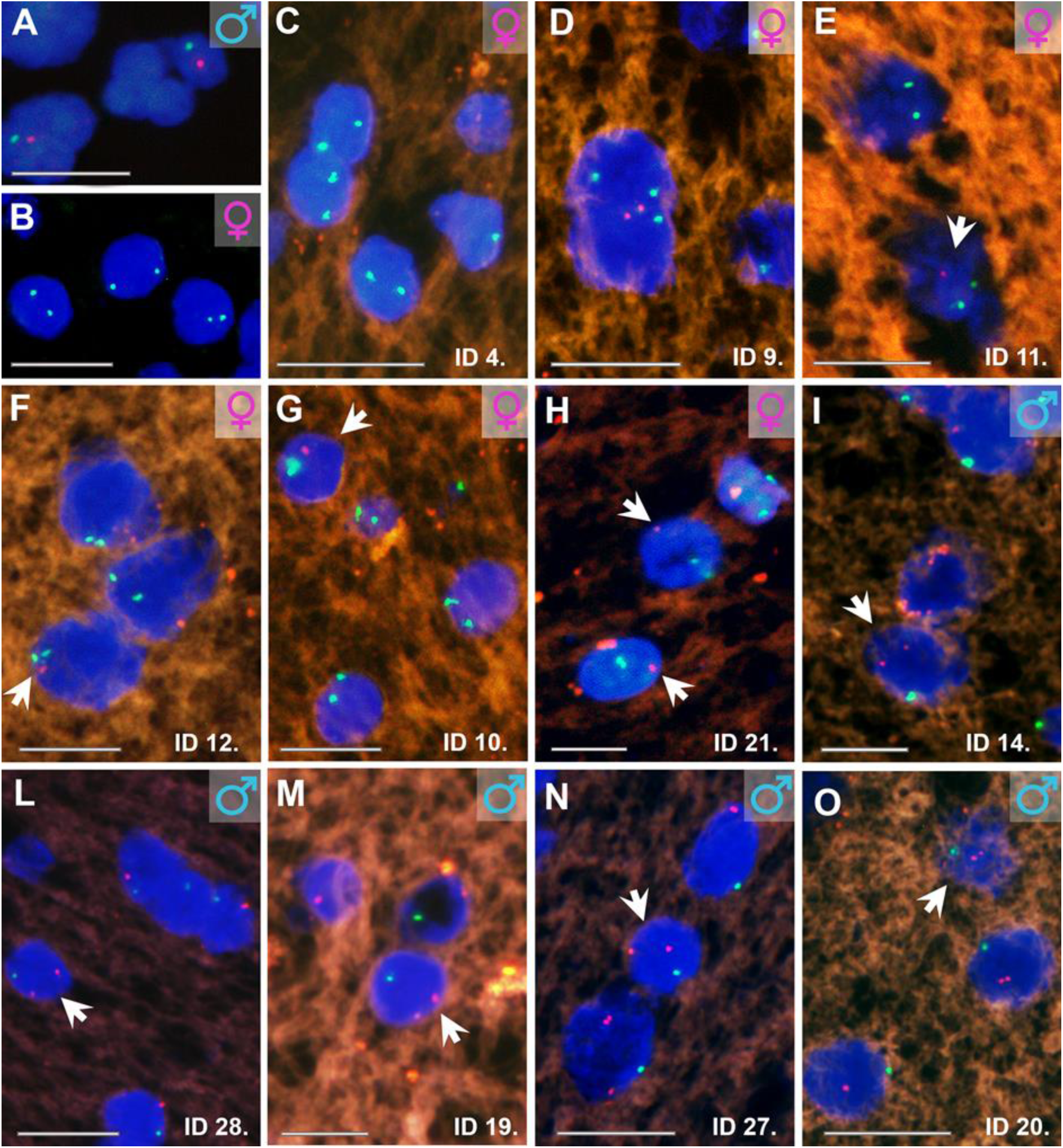
Fluorescence in situ hybridisation (FISH) for centromeric X and Y probes in control and MOGHE brain tissue samples. **Legend Figure 3.** Panels (**A–O**) show representative nuclei counterstained with DAPI (blue) and hybridised with dual-colour centromeric probes: X (green) and Y (red). (**A**) Control male sample from an individual with TLE, with one green and one red signal. (**B**) Control female sample from an individual with TLE with two green signals and no red signal. (**C**) Female individual with MOGHE and negative for Y-chromosome gain. The nuclei display two green X-signals and an additional red Y-signal, indicating Y-chromosome gain in otherwise XX cells. (**D–H**) Female individuals with MOGHE and positive for Y-chromosome gain (IDs 9, 11, 12, 10, 21). Some nuclei display two green X-signals and an additional red Y-signal, indicating Y-chromosome gain in otherwise XX cells. (**I–O**) Male individuals with MOGHE (IDs 14, 28, 19, 27, 20), showing nuclei with one green signal and two red signals, consistent with Y-chromosome gain. Scale bar in A, B, C, G, O = 10 µm, scale bar in D, E, F, H, I, L, M, N = 5 µm.

**Figure 4.**
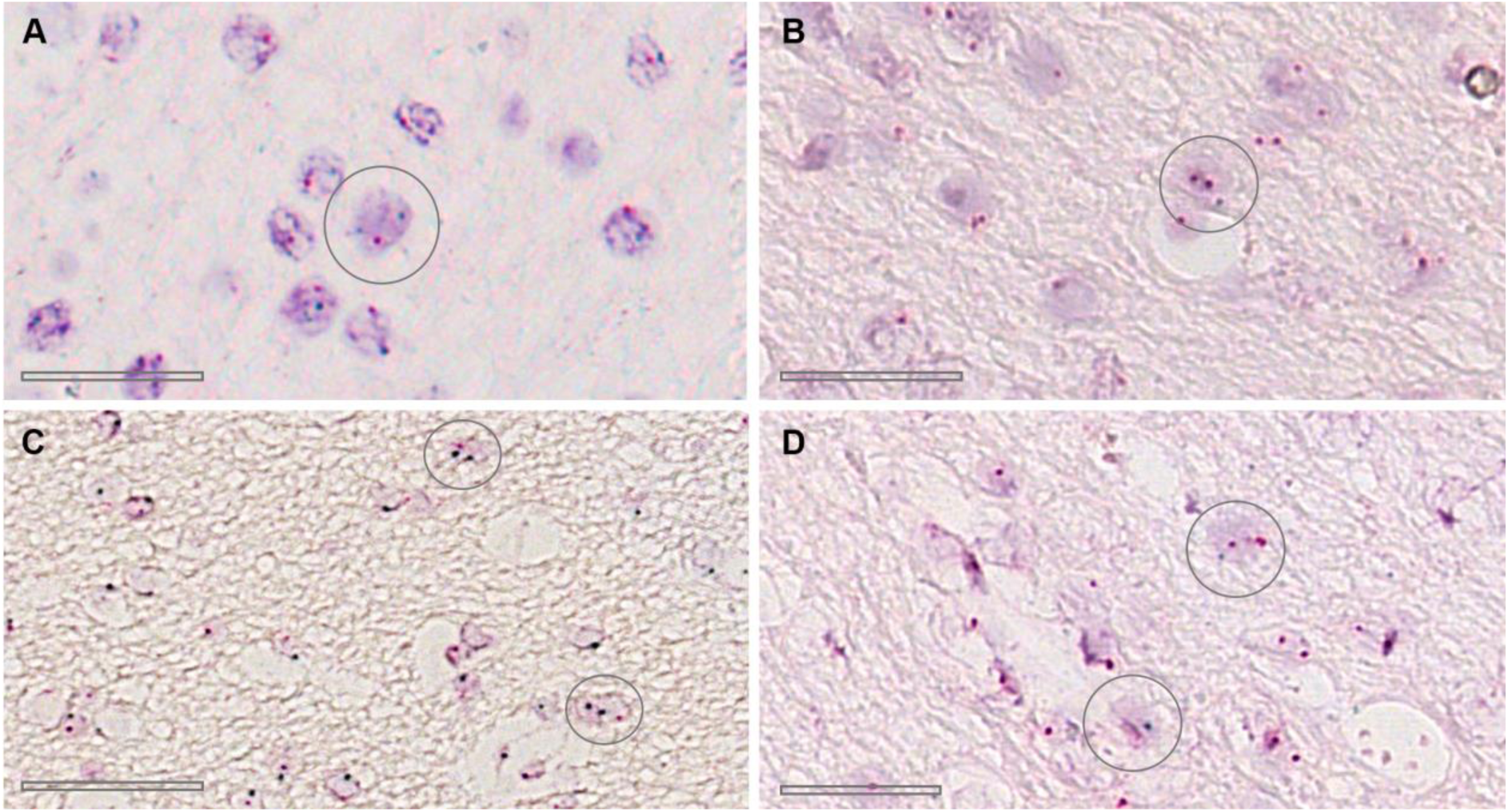
Chromogenic in situ hybridisation (CISH) for centromeric X and Y probes in MOGHE brain tissue samples with Y chromosome gains. **Legend Figure 4**. Panels show representative nuclei in lesional white matter, with the X chromosome visualised in red and the Y chromosome visualised in green. (**A**) Male MOGHE case (ID 17) with a nucleus containing one X and two Y chromosomes. (**B**) Female MOGHE case (ID 21) with a nucleus containing two X and one Y chromosome. (**C**) Male MOGHE case (ID 15) with a nucleus containing one X and two Y chromosomes. (**D**) Female MOGHE case (ID 11) with two nuclei, each containing two X and one Y chromosome. Highlighted circles indicate representative nuclei. Scale bar in A, B, D = 25 µm, scale bar in C= 50µm. Note that the probe colour scheme for CISH is opposite to that for FISH (see Figure 2).

### PCR validation targeting *SRY* confirms the presence of Y-chromosome sequences in brain tissue with MOGHE from Female individuals

PCR targeting the SRY locus was performed in DNA extracts from seven female patients (Table 1). This subset included four females with clearly positive Y-chromosome gain in CNV plots, one female with ambiguous or intermediate CNV results, and two females with CNV plots indicating absence of Y-chromosome gain (ID 4 and 5), which served as negative controls. Additionally, the DNA from a female patient with a Ganglioglioma was included as a negative control, and DNA from a male patient with temporal lobe epilepsy (TLE) served as a positive control (**Figure 5**). DNA was extracted separately from the lesional white matter and the adjacent overlying cortex, guided by manual microdissection using histological and OLIG2 immunostaining to identify regions of increased oligodendroglial density (**Fig. 5A**). Y-specific amplicons were detected in all females with CNV-confirmed Y-chromosome gains, as well as in the case with an intermediate log2 ratio value (ID 9) or an inconclusive CNV plot (ID 10), thereby confirming the presence of Y-chromosomal sequences even when CNV data were unclear. No amplification was observed in the CNV-negative controls (ID 4 and 5) or in the negative control. Within each individual, Y-specific amplicons were consistently observed in the lesional white matter but absent in the paired cortical tissue, highlighting the spatially restricted distribution of Y-positive cells in areas of oligodendroglial hyperplasia (**Fig. 5B**).

**Figure 5.**
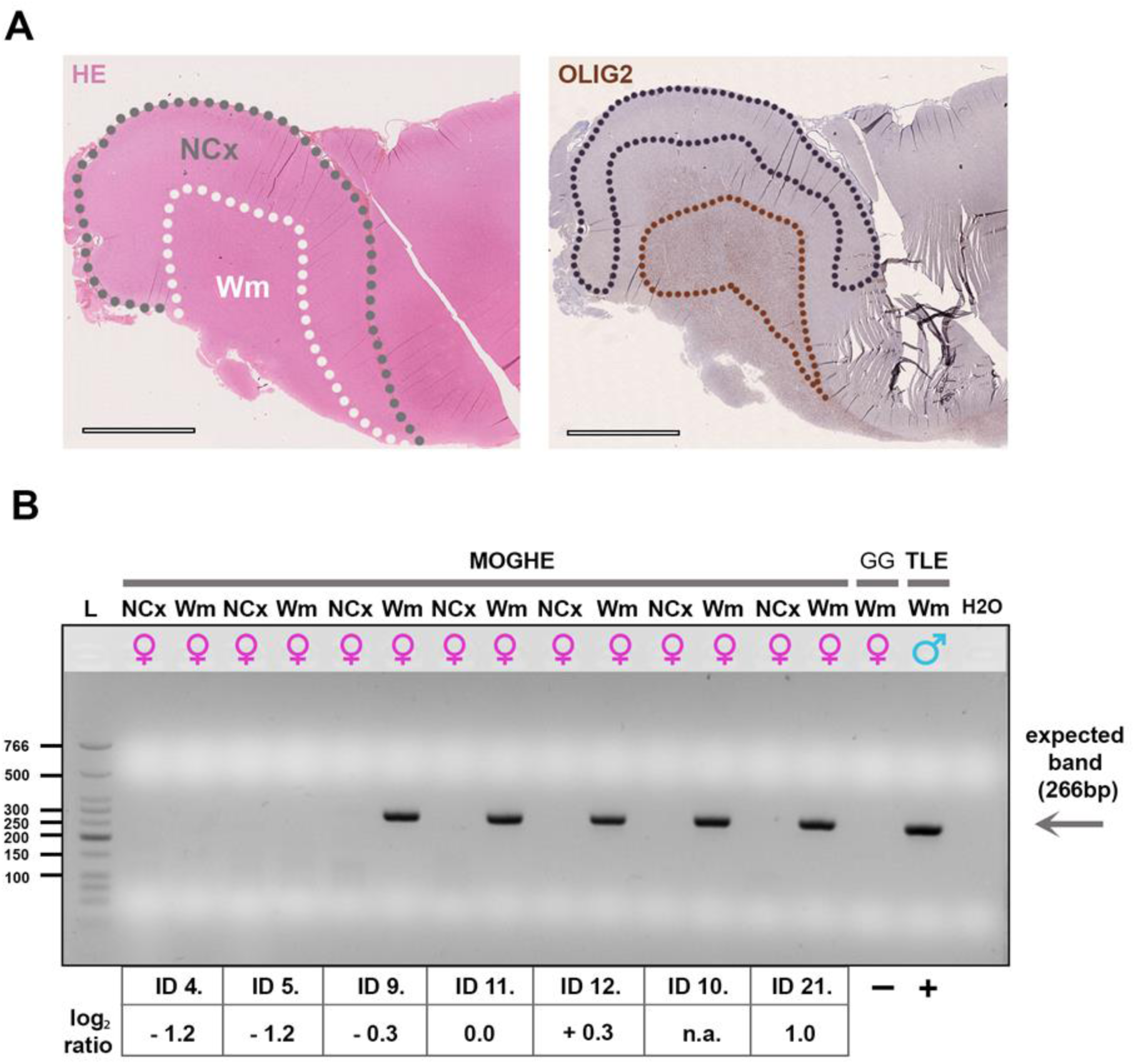
PCR amplification of the Y chromosome–specific SRY locus in female MOGHE patients and controls. **Legend Figure 5.** (**A**) Representative histological images illustrating the regions selected for microdissection prior to PCR analysis. The hematoxylin and eosin (HE) staining well distinguishes between cortical (NCx) and subcortical white matter (WM) in MOGHE. The adjacent OLIG2 immunostain highlights the increased density of oligodendroglia. Dotted lines mark examples of manually micro-dissected regions. (**B**) Y-specific amplicons of a 266 bp fragment were consistently detected in the white matter lesions but absent in the paired cortical tissue, demonstrating a lesion-restricted distribution of Y-positive cells. The 266 bp SRY amplicon was observed in all females with CNV log2 ratios ≥ 0.0 and in the male positive control (TLE). DNA from a female patient with Ganglioglioma (GG) was included as a negative control. L indicates the DNA size marker (ladder; sizes shown on the left).

The presence of *SRY*-specific amplicons in lesional tissue, but not in matched non-lesional cortex, provided orthogonal validation of the CNV analyses and in situ hybridisation findings and demonstrated lesion-restricted Y chromosome gain mosaicism. Conversely, none of the CNV-negative cases yielded positive ISH or PCR results. These results corroborated the CNV and *in situ* hybridisation data, confirming that Y-chromosome gains are spatially confined to areas of oligodendroglial hyperplasia within the MOGHE lesions.

## Discussion

This study identifies recurrent gains of Y-chromosomal sequences in 72% of surgically resected brain tissue samples obtained from 29 individuals with histopathologically confirmed MOGHE, in 16 males (*n*=19, 84.2%) and five females (*n*=10, 50%). Using different approaches of DNA methylation–derived CNV analysis, CISH, FISH, and PCR targeting the *SRY* locus, we demonstrate that Y-chromosome gains were spatially confined to lesional white matter and were specifically enriched in clusters of oligodendroglial hyperplasia. These results provide a genetic cause in all our individuals with MOGHE, either by X-linked *SLC35A2* alterations (n=8, 28%), a gain of the Y chromosome (n=9, 31%), or both conditions together (n=12, 41%), suggesting that MOGHE is a sex chromosome-linked disease entity.

Whilst all MOGHE samples showed the same histopathology phenotype with respect to oligodendroglial cell densities, heterotopic neurons in the white matter, or cell proliferation, molecular-genetic MOGHE subtypes revealed clinically relevant differences in disease onset and lesional volumes. The MOGHE subgroup carrying *SLC35A2* and Y-chromosome alterations was numerically the biggest (41% of the entire cohort), had the largest lesion volume, often expanding from the frontal lobe into posterior regions or affecting the entire hemisphere, and had the earliest disease onset. MOGHE lesions of patients carrying Y-chromosome alterations alone were also large, and presented with earlier seizure onset than the *SLC35A2* variant subgroup. However, these observations were obtained from a small patient cohort of 29 individuals and did not reach statistical significance. Independent cohort studies will thus be necessary to confirm and validate our findings.

The detection of Y-chromosomal material in females demonstrates the presence of sex chromosome mosaicism or microchimerism in MOGHE. Somatic sex chromosome mosaicism is increasingly recognized as a spectrum that extends beyond classical germline aneuploidies in Turner syndrome, where Y-chromosome-positive cell populations have been documented in otherwise phenotypic females ^49^. Similarly, male microchimerism has been observed in human brain tissue, likely originating from foetal–maternal cell exchange ^50^, and could provide an alternative explanation for the presence of Y-positive cells in female patients.

The occurrence of Y-chromosome gains in females may reflect abnormal chromosomal segregation during mitosis in early embryonic development, resulting in somatic mosaicism. Alternatively, male microchimerism is a plausible explanation: male-derived cells can persist in maternal tissues following pregnancy with a male fetus and have been detected in multiple organs, including the brain ^50,51^. Whether Y-positive cells in female MOGHE patients extend beyond the brain remains unknown and will require examination of extracerebral tissues. In males, Y-chromosome gains are most likely attributable to mitotic errors during early neurodevelopment, leading to somatic mosaicism.

A striking feature of our study is the lesion-restricted distribution of Y-positive nuclei. One explanation is clonal expansion of Y-positive cells within the pathological microenvironment, which renders them detectable in lesions but not in the surrounding cortex. Alternatively, chronic inflammation, oxidative stress, and vascular changes characteristic of lesional tissue could increase blood–brain barrier permeability, facilitating selective infiltration or persistence of Y-positive cells ^52,53^. Another possibility is that Y-chromosome gains provide a proliferative or survival advantage to oligodendroglial precursors in the diseased microenvironment, thereby driving their enrichment in lesional tissue ^54^.

Y-chromosome gains were identified in both *SLC35A2*-mutant and wild-type patients, indicating that this alteration represents an independent molecular feature of MOGHE. The restriction of Y-positive nuclei to lesional oligodendroglial clusters suggests a potential pathogenic role, either by altering the proliferative or plasticity of oligodendroglia or by perturbing the local microcircuitry. While the specific contribution of Y-linked genes remains to be determined in this disease condition, dosage effects on genes involved in neurodevelopment, synaptic signalling, or myelination could also influence epileptogenesis. These observations align with broader evidence that both X- and Y-chromosome abnormalities can impact brain development and increase seizure susceptibility ^25,55–58^.

Importantly, the combined use of CNV analysis, FISH, and PCR provided complementary strengths and increased diagnostic confidence. While DNA methylation–derived CNV profiles captured Y-chromosome gains in the majority of patients, individual cases with inconclusive or ambiguous CNV results could be resolved by FISH or PCR. These findings underscore that orthogonal validation by in situ hybridisation or PCR not only supports the robustness of CNV-based approaches but also provides an essential means of resolving borderline cases.

Several limitations must be acknowledged. The present study cannot definitively distinguish between somatic mosaicism and microchimerism as the source of Y-positive cells. Genomic approaches at single-cell resolution will be required to resolve the structural nature of Y-chromosome gains and their cellular lineage ^59^. In addition, further studies are necessary to establish whether these alterations directly contribute to the pathogenesis of MOGHE or if they are just an associated finding without a direct effect.

In conclusion, we provide convergent evidence that Y-chromosome gains represent a recurrent and lesion-restricted feature of MOGHE. The high incidence of Y-chromosomal material in both male and female patients for this type of focal epilepsy underscores the need to further investigate the contribution of sex chromosome biology to the pathogenesis of this increasingly recognised focal malformation.

## Fundings

IB, KK, LH, KrK, and EC were supported by the Deutsche Forschungsgemeinschaft (DFG, German Research Foundation) project number 460333672–CRC1540 Exploring Brain Mechanics. Confocal microscopy was enabled on a Leica Stellaris 8 laser scanning microscope, funded by Deutsche Forschungsgemeinschaft (DFG, German Research Foundation)—project 441730715.

## Competing interests

The authors report no competing interests.

## Data availability

The datasets generated and analysed during the current study are not publicly available due to patient privacy and ethical restrictions, but are available from the corresponding author upon reasonable request.

## Data Availability

All data produced in the present work are contained in the manuscript

## Notes

### Competing Interest Statement

The authors have declared no competing interest.

### Funding Statement

This study was funded by: IB, KK, LH, KrK, and EC were supported by the Deutsche Forschungsgemeinschaft (DFG, German Research Foundation) project number 460333672-CRC1540 Exploring Brain Mechanics. Confocal microscopy was enabled on a Leica Stellaris 8 laser scanning microscope, funded by Deutsche Forschungsgemeinschaft (DFG, German Research Foundation) project 441730715.

### Author Declarations

Medical Faculty of the Friedrich-Alexander University Erlangen-Nuremberg (#193_18B, 18-192_1-Bio) gave ethical approval to this work University of Muenster (2015-088-f-S) gave ethical approval to this work

